# Normative volumetric growth modeling of the whole fetal body, placenta and amniotic fluid for 3-dimensional T2-weighted magnetic resonance imaging

**DOI:** 10.64898/2026.01.16.26344259

**Authors:** Alena Uus, Megan Hall, Charline Bradshaw, Anangsha Kumar, Jordina Aviles Verdera, Sara Neves Silva, Aysha Luis, Hadi Waheed, Pedro Alarcon Gil, Lucilio Cordero-Grande, Jacqueline Matthew, Vanessa Kyriakopoulou, Maria Deprez, Kathleen Colford, Alexia Egloff Collado, Joseph V. Hajnal, Mary Rutherford, Jana Hutter, Lisa Story

**Affiliations:** Research Department of Early Life Imaging, School of Biomedical Engineering and Imaging Sciences, King’s College London, London, UK; Department of Women and Children’s Health, King’s College London, London, UK; Biomedical Image Technologies, ETSI Telecomunicacion, Universidad Politécnica de Madrid and CIBER-BBN, Madrid, Spain; Research Department of Imaging Physics and Engineering, School of Biomedical Engineering and Imaging Sciences, King’s College London, London, UK; Smart Imaging Lab, Radiological Institute, University Hospital Erlangen, Erlangen, Germany; Fetal Medicine Unit, Guy’s and St Thomas’ NHS Foundation Trust, London, UK

**Keywords:** fetus, amniotic fluid, placenta, volumetry, magnetic resonance imaging, normative modeling

## Abstract

**Background:** Volumetric assessment of the fetus, placenta and amniotic fluid is clinically valuable, but MRI volumetry is rarely performed in clinical practice because of the required labour-intensive manual segmentation of motion-corrupted 2-dimensional (2-D) stacks. Existing deep-learning approaches typically segment single structures in 2-D motion-corrupted stacks, are, however limited in accuracy by slice misalignment. No current method provides a reliable automated solution for whole-uterus volumetry in 3-D reconstructed MRI. Furthermore, normative ranges for computation of centiles are currently missing.

**Objective:** To develop an automated pipeline for whole-uterus volumetry in 3-D T2-weighted fetal MRI and to generate normative growth models for fetal, placental and amniotic fluid volumes in healthy pregnancies with confirmed delivery at term.

**Materials and methods:** Deformable slice-to-volume 3-D reconstruction was applied to motion-corrupted T2-weighted (T2W) stacks from 0.55T-3T MRI, and a 3-D UNet was trained to segment fetus, placenta and amniotic fluid on the resulting reconstructed 3-D images. A reporting tool generates centiles, z-scores and structured HTML outputs. Automated segmentation was performed in 357 healthy control datasets from 16-41 weeks gestational age (GA) range with confirmed delivery at term. After visual checks of segmeted labels and minor refinements, GA-based quadratic normative volumetry models were derived and correlations with maternal and fetal characteristics assessed. The utility of the pipeline for clinical research was further evaluated using 95 longitudinal scans from 42 fetuses and 86 preterm (≤ 32 weeks at delivery) pregnancies.

**Results:** Automated segmentation produced accurate 3-D labels, with only small local corrections (< 1% volume difference) required in the control cohort(< 25% of the datasets). Fetal and placental volumes increased across gestation, while amniotic fluid volume peaked mid-pregnancy and declined toward term. Volumes and centiles correlated with maternal size and birth weight. Longitudinal scans showed individual fetal and placental trajectories closely following the normative curves, with greater variability in amniotic fluid. Preterm pregnancies showed significantly lower fetal, placental and amniotic fluid volumes and centiles than the controls with confirmed delivery at term.

**Conclusion:** This study introduces an automated whole-uterus volumetry pipeline and corresponding normative 3-D MRI growth models. The method provides robust, standardised volumetric assessment of fetal, placental and amniotic fluid development and offers a practical tool for evaluating growth patterns in both normal and high-risk pregnancies.

## Introduction

Fetal MRI is a valuable adjunct to antenatal ultrasound for assessing fetal and placental anatomy, growth and function. Compared to ultrasound, MRI offers superior soft-tissue contrast, a wider field of view encompassing the entire uterus, and true three-dimensional (3-D) information, improving anatomical assessment and diagnostic confidence [1]. This makes MRI well suited for quantifying volumes of key intrauterine structures—the fetus, placenta and amniotic fluid, serving as objective markers of fetal growth, placental sufficiency and overall pregnancy health. These volumes are conventionally obtained from segmentations of MRI stacks.

Quantitative global intrauterine volumetry is clinically informative and important for multiple reasons. Fetal body volume reflects overall fetal size and, when plotted against gestational-age-adjusted centiles, helps identify fetuses that are unusually small or large for gestation, which may indicate fetal growth restriction or macrosomia [2] conditions for which early detection is critical for timely clinical intervention. MRI-derived total fetal volume is also widely used as a denominator for volumetry of individual fetal organs (e.g., brain, lungs) [3–5] and for quantitative flow estimation in fetal cardiac MRI [6], making accurate whole-fetus segmentation essential for many quantitative MRI applications.

Placental volume provides a macroscopic indicator of placental development and functional capacity. Smaller placental volumes are associated with placental insufficiency and conditions such as fetal growth restriction, preeclampsia and hypertensive disorders, reflecting impaired villous development and reduced fetal growth [7]. Larger placental volumes may occur in gestational diabetes or anemia due to increased metabolic demand [7]. Placental size also correlates strongly with birth weight and neonatal outcomes [8], supporting its value as a complementary biomarker alongside Doppler Ultrasound.

Amniotic fluid volume reflects fetal renal and gastrointestinal function, placental health and membrane integrity [9, 10]. Oligohydramnios (low amount of amniotic fluid) may result from placental insufficiency or fetal growth restriction through reduced renal perfusion, and is also characteristic of PPROM, where sustained fluid loss can additionally impact lung development. Polyhydramnios (excess of amniotic fluid) is commonly associated with maternal diabetes or fetal gastrointestinal obstruction (e.g., tracheoesophageal fistula) and is linked to increased perinatal risk. As amniotic fluid is influenced by multiple fetal and placental pathways, its volumetric assessment provides a useful integrated indicator of intrauterine health.

MRI-based 3-D volumetry provides reproducible, objective and quantitative measurements that overcome the limitations of subjective interpretation, isolated 2-D metrics and inherently suboptimal formula-based weight estimates commonly used in ultrasound. Despite these advantages, MRI-based volumetry is rarely performed in routine practice. Most research studies continue to rely on labour-intensive manual segmentation of motion-corrupted balanced steady-state free precession (bSSFP) or T2-weighted single-shot turbo spin echo (SSTSE) stacks [11–15]. This approach is slow, operator dependent and error prone, particularly for large structures such as the amniotic cavity, fetal body or placenta. Recent deep-learning methods offer partial automation, yet most are trained directly on raw motion-corrupted stacks and typically target a single region of interest such as the fetal body [5, 16], placenta [17, 18] or amniotic fluid [19]. Multi-structure approaches [20, 21] remain limited by narrow coverage of gestational-age, small training cohorts, and single-protocol datasets. Crucially, slice misalignment and through-plane motion in raw stacks restrict volumetric accuracy, and no existing method enables reliable simultaneous volumetry of fetus, placenta and amniotic fluid in true 3-D images.

There are also no combined normative growth charts for these three compartments derived specifically from healthy control pregnancies across multiple field strengths. Given the physiological interdependence of fetal, placental and amniotic volumes, combined assessment has been shown to improve diagnostic certainty [22]. Existing MRI volumetry studies are however heterogeneous, typically focusing on a single structure, restricted GA ranges or mixed referral populations. Only one recent study reports a normative model for MRI-derived fetal body volume based on 260 healthy fetuses (16-36 weeks GA) [15]. Comparable normative datasets are lacking for MRI-derived placental or amniotic fluid volumes. Consequently, clinicians lack MRI-based centiles and z-scores to determine whether measured volumes fall within expected ranges or to evaluate how the three compartments relate to one another. Establishing normative curves is therefore essential a robust standardisation framework against which individual measurements can be interpreted, which is central to the clinical utility of global intrauterine volumetry. Characterising cross-compartment and longitudinal relationships then enables early identification of abnormal developmental trajectories. Deformable slice-to-volume registration (DSVR) [23] method addresses these challenges by reconstructing high-resolution isotropic 3-D images from multiple motion-corrupted T2W stacks. This process removes inter-slice misalignment and restores anatomical continuity, enabling more reliable segmentation and volumetry [24]. However, existing 3-D fetal MRI segmentation efforts have focused primarily on the fetal brain or individual fetal organs, and no prior work has provided a solution for combined whole-uterus volumetry—including the fetus, placenta and amniotic fluid—in reconstructed 3-D MRI

### Contributions

This work fills key gaps in MRI-based volumetric assessment of the whole uterus by introducing the first integrated pipeline for automated fetal, placental and amniotic fluid volumetry in motion-corrected 3-D reconstructed T2-weighted fetal MRI across 0.55-3T field strengths. The pipeline provides:(1) automated deep-learning segmentation of all three structures in 3-D reconstructed volumes; (2) normative growth models derived from 357 confirmed healthy term-control scans covering 16-41 weeks gestational age; and (3) a reporting tool that computes centiles and z-scores and generates a structured report for clinical interpretation. We further demonstrate the utility of this framework by analysing patient-specific volumetric trajectories in 95 longitudinal scans from 42 fetuses, and by evaluating deviations in an abnormal preterm birth cohort (n = 86).

## Materials and methods

### Cohort, image acquisition and preprocessing

The study uses fetal MRI datasets acquired at St Thomas’ Hospital, London as part of five ethically approved studies (REC 16/LO/1573, 21/SS/0082, 21/LO/0742, 22/YH/0210, 14/LO/1806, 19/LO/0736). All imaging and data handling procedures adhered to relevant ethical and clinical governance standards, and written informed consent was obtained from all participants.

The T2W SSTSE sequences were acquired across three MRI systems using protocol variations representative of clinical practice:

- 3T Philips Achieva (n = 241; Philips Healthcare, The Netherlands), 32-channel cardiac coil; TE = 180 ms; in-plane resolution 1.25 × 1.25 mm; slice thickness 2.5 mm; −1.5 mm slice gap; 5-6 stacks.
- 1.5T Philips Ingenia (n = 122; Philips Healthcare, The Netherlands), 28-channel torso coil; TE = 80/180 ms; in-plane resolution 1.25 × 1.25 mm; slice thickness 2.5 mm; −1.25 mm slice gap; 6-9 stacks.
- 0.55T Siemens MAGNETOM Free.Max (n = 143; Siemens Healthineers, Germany), 6-element flexible + 9-element spine coil; TE = 105-106 ms; in-plane resolution 1.48 × 1.48 mm; slice thickness 4.5 mm; 9-12 stacks [25].

The complete dataset (training + analysis) comprises 506 scans from 16-41 weeks’ gestation from the following groups:

- 357 term controls, with confirmed delivery ≥ 37 weeks and no reported fetal, placental or maternal abnormalities.
- 86 high-risk cases, consisting of pregnancies with confirmed preterm delivery ≤ 32 weeks or termination following PPROM.
- 63 additional datasets used exclusively for model training, including cases with various fetal and maternal anomalies, unknown pregnancy outcomes or outside the main inclusion criteria.

For each case, whole-uterus 3-D reconstruction was performed using deformable slice-to-volume reconstruction (DSVR) [23] implemented in the SVRTK framework (https://github.com/SVRTK/auto-proc-svrtk). This included DL-based masking of the uterus, semi-automated template stack quality control and reconstruction to 1.2 mm isotropic resolution. An additional 28 datasets were reconstructed using a more recent deep generative prior DSVR method [26].

General inclusion criteria for volumetric analysis were: (1) singleton pregnancy; (2) acceptable quality 3-D reconstruction [24] with complete uterine coverage; (3) sufficient visibility of fetal body, placenta and amniotic fluid.

The normal control cohort consists of pregnancies that resulted in healthy term delivery without reported fetal, placental or maternal structural anomalies. The preterm cohort represents early delivery (≤32 weeks), including cases complicated by PPROM, and therefore reflects clinically relevant intra-uterine pathology. In addition to pregnancy outcomes, we collected available maternal demographic information, including ethnicity, weight and height, when reported. Birth weight centiles were calculated using neonatal population charts from the Fetal Medicine Foundation [27].

### Automated segmentation

We defined a parcellation protocol for global intrauterine anatomy, comprising four labels: whole fetus (further separated into fetal head and fetal body), placenta, amniotic fluid, and the umbilical cord is set to zero label, which is excluded from volumetry.

For automated segmentation (Fig 1), we trained a deep-learning model based on a classical 3D U-Net architecture [28], implemented in the MONAI framework [29]. Training was performed using 223 manually curated 3D DSVR reconstructions, spanning 17-39 weeks’ gestation and acquired across 0.55T, 1.5T, and 3T MRI systems. The training cohort included normal controls with confirmed delivery at term, high-risk PPROM preterm cases, and fetuses with a range of structural abnormalities, ensuring robustness in diverse anatomy and imaging conditions.

**Fig. 1:**
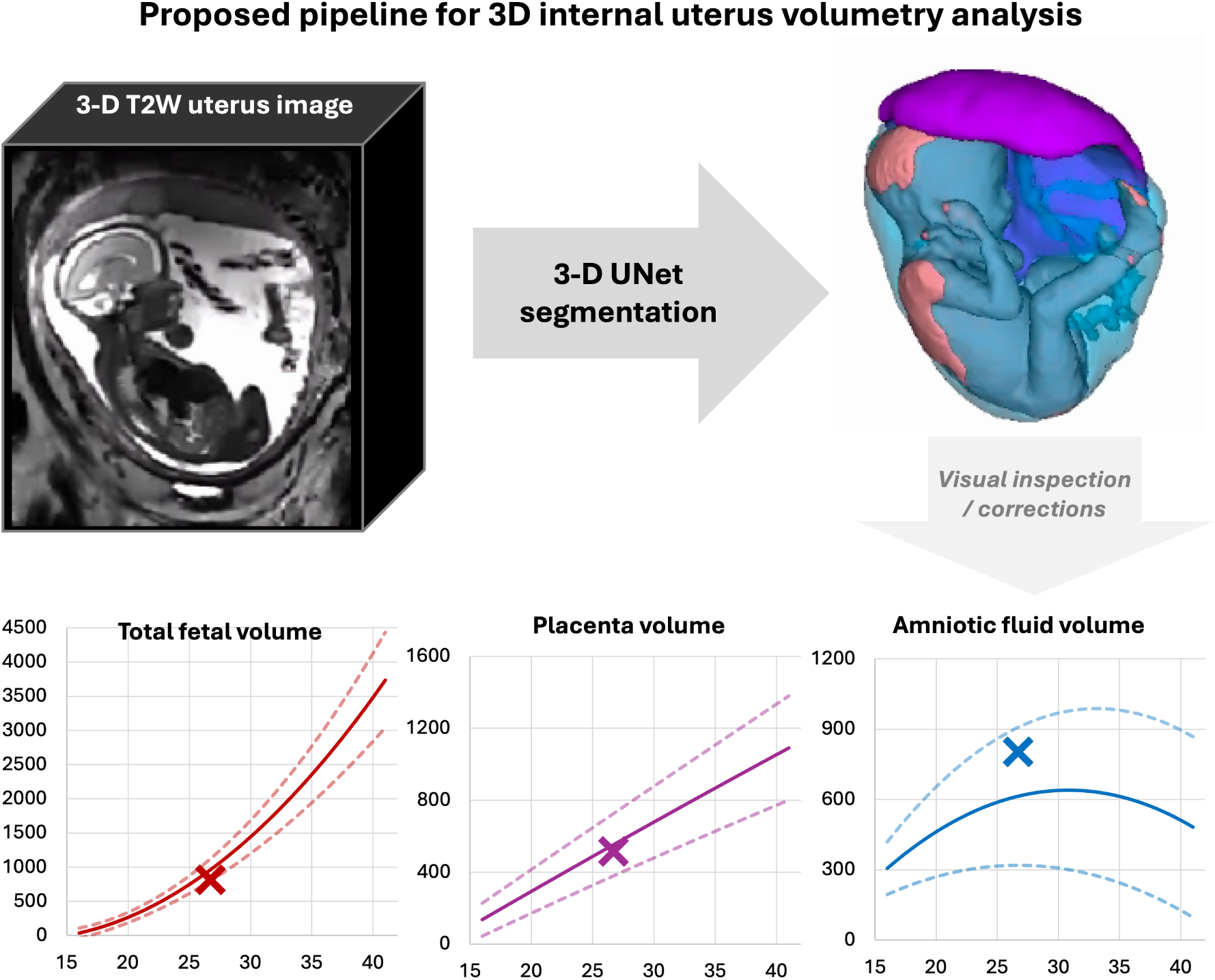
Proposed pipeline for automated volumetric analysis for 3-D T2W fetal MRI. *DSVR* deformable slice-to-volume reconstruction, *3-D* 3-dimensional, *T2W* T2-weighted

Ground-truth labels were generated through an iterative process combining existing in-house DL models, manual segmentations in ITK-SNAP [30], and multiple rounds of interleaved model training and manual refinement to improve anatomical consistency and boundary accuracy.

Model performance was evaluated on an independent test set of 43 datasets (0.55T, 1.5T, 3T) not used during training. Evaluation included (1) comparison with manually refined automated labels and (2) qualitative quality scoring for each anatomical region of interest (ROI), assessing delineation of fetal body, placenta, and amniotic fluid across the range of gestational ages and field strengths.

### Normative growth modeling

The trained model was applied to 357 normal-control 3D DSVR T2-weighted whole-uterus reconstructions acquired across 0.55T, 1.5T, and 3T systems. All automated labels were visually inspected and, where necessary, manually refined in ITK-SNAP [30] to ensure accurate delineation of the fetus, placenta, and amniotic fluid.

For each case, volumetric measurements (in cubic centimetres) were extracted from the corrected labels. These volumes were then used to construct normative growth models for fetal, placental, and amniotic fluid volumes across gestation. Gestational-age trajectories for the 50th, 5th, and 95th centiles were estimated using quadratic regression following established methodology [31].

Statistical analysis in the normal-control cohort was performed using analysis of covariance (ANCOVA) implemented in the statsmodels Python library, allowing assessment of associations between volumes, gestational age, and maternal or fetal characteristics.

The resulting normative models were incorporated into a centile calculator (Excel format) and a Python-based reporting tool that generates structured HTML reports including segmentation visualisation and centile plots. In addition, estimated fetal weight was computed from fetal volume based on formula defined by Baker et al. (*EFW_Baker_*(*kg*) = 1.031*·V_fetus_* +0.12) [32]. All automated segmentation components, centile calculators, and reporting scripts are publicly available online at the auto-SVRTK repository (https://github.com/SVRTK/auto-proc-svrtk).

### Volumetric analysis of term vs. preterm cohorts

To assess the clinical utility of the segmentation pipeline and normative models, we performed a comparative analysis between the 86 preterm cases (delivery ≤ 32 weeks, including PPROM) and the normal-control cohort. All preterm datasets were segmented using the same automated network and underwent visual quality review, with manual refinements applied when necessary to ensure consistency across groups.

Volumetric comparisons were conducted using both absolute volumes and gestational-age-adjusted centiles, allowing evaluation of whether fetuses destined for preterm birth deviated from expected growth patterns at the time of MRI. Statistical analysis was performed using ANCOVA, correcting for gestational age at MRI.

## Results

### Automated segmentation

The trained segmentation model was evaluated on 43 independent datasets spanning all three field strengths, a wide gestational-age range, varied fetal and placental positions, and both term and preterm pregnancies (Fig 2a). Example segmentations at 17-38 weeks’ gestation are shown in Fig 3. Across this range, the 3-D labels showed smooth, anatomically plausible boundaries and realistic appearances for all structures.

**Fig. 2:**
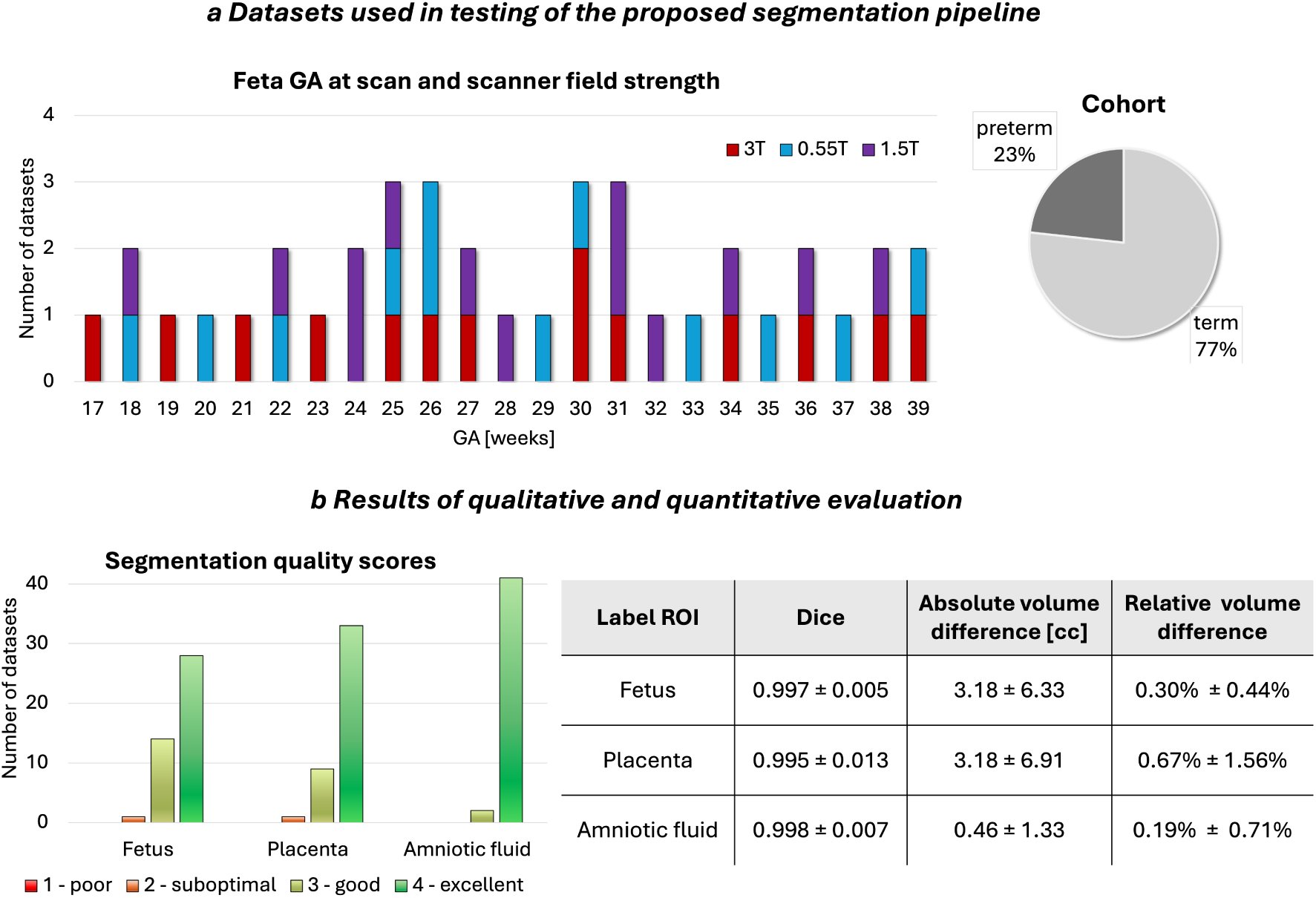
a) Exemplary fetal MRI datasets used in testing of the proposed segmentation pipeline. b) Qualitative and quantitative results of testing. *GA* gestational age, *ROI* region of interest, *T* Tesla

**Fig. 3:**
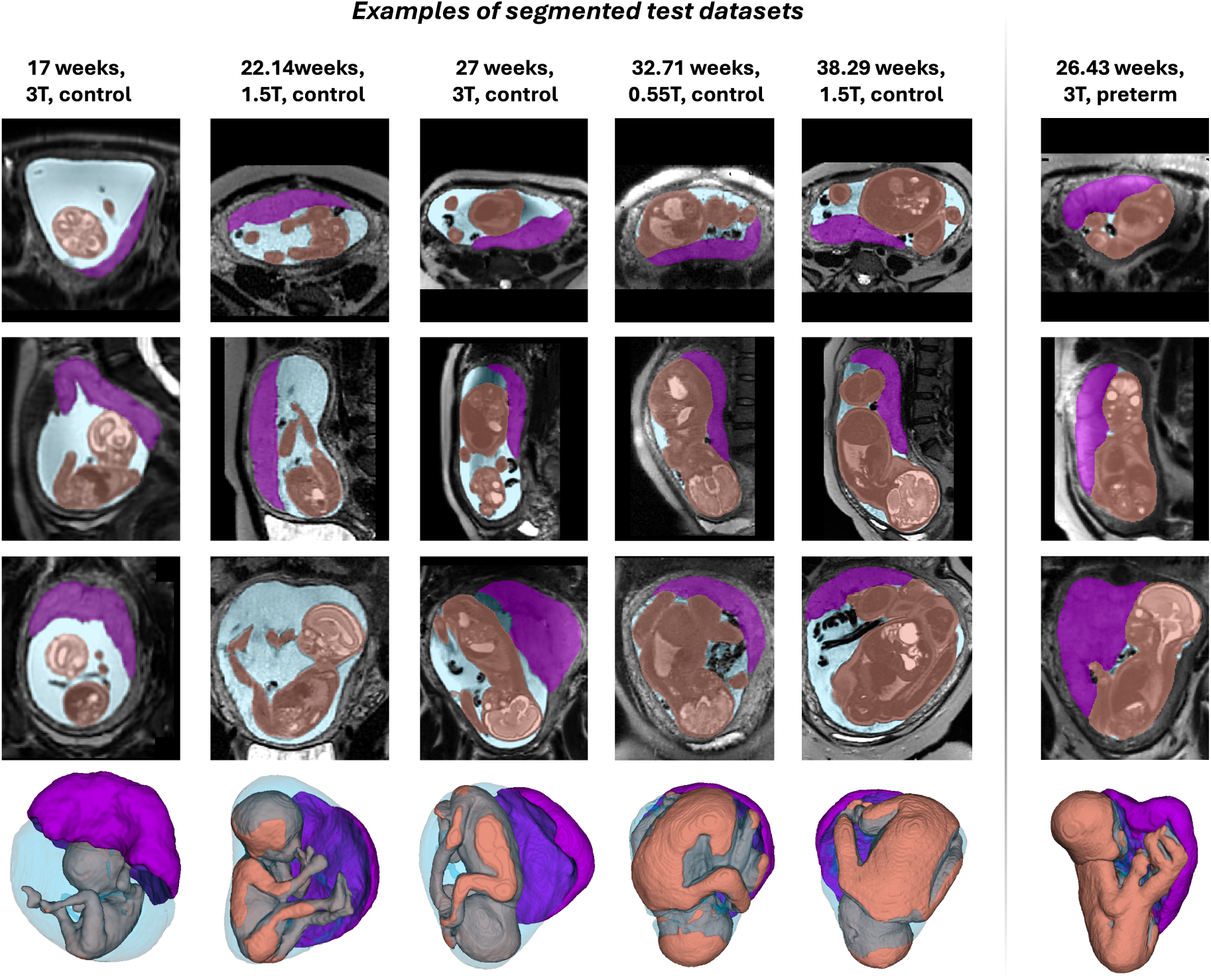
Examples of 3-D segmentations for test cases from different gestational ages, scanner field strengths and cohorts. *GA* gestational age, *T* Tesla

Each segmentation was inspected in both 2-D slices and 3-D renderings, and quality was graded on a 1-4 scale (1 = poor, 2 = suboptimal, 3 = good, 4 = excellent) for every structure. A fetal MRI researcher then manually refined each case to create a reference label, enabling comparison between automated and refined segmentations using Dice score and absolute/relative volume differences. The qualitative scores and quantitative results are summarised in Fig 2b.

Overall, segmentation quality was high, with most structures receiving good or excellent ratings. The average QC scores were 3.63 *±* 0.54 for the fetus 3.74 *±* 0.49 for placenta, and 3.95 *±* 0.21 for amniotic fluid. Only one fetal and one placental segmentation were scored as suboptimal. This was consistent with the quantitative metrics, which showed high Dice similarity and low relative volumetric error. Most refinements were required at the placenta-myometrium interface, where contrast can be limited and placental morphology irregular, and around fetal limbs, which may be difficult to distinguish from the umbilical cord - particularly in PPROM cases with reduced amniotic fluid and in 0.55T datasets where image quality is lower.

### Normative growth modeling

Summary characteristics of the normal control cohort are shown in Fig 4. Automated segmentation outputs for all 357 control datasets were visually reviewed. Most were immediately suitable for volumetric analysis; minor manual refinement was required in 87 cases (< 25% of all datasets), predominantly at the placenta-myometrium interface and around fetal limbs, where contrast is limited. These refinements were fast (typically < 2 minutes per case) and had no meaningful impact on the final growth models. Z-scores and centiles were subsequently computed for all volumetric measures.

**Fig. 4:**
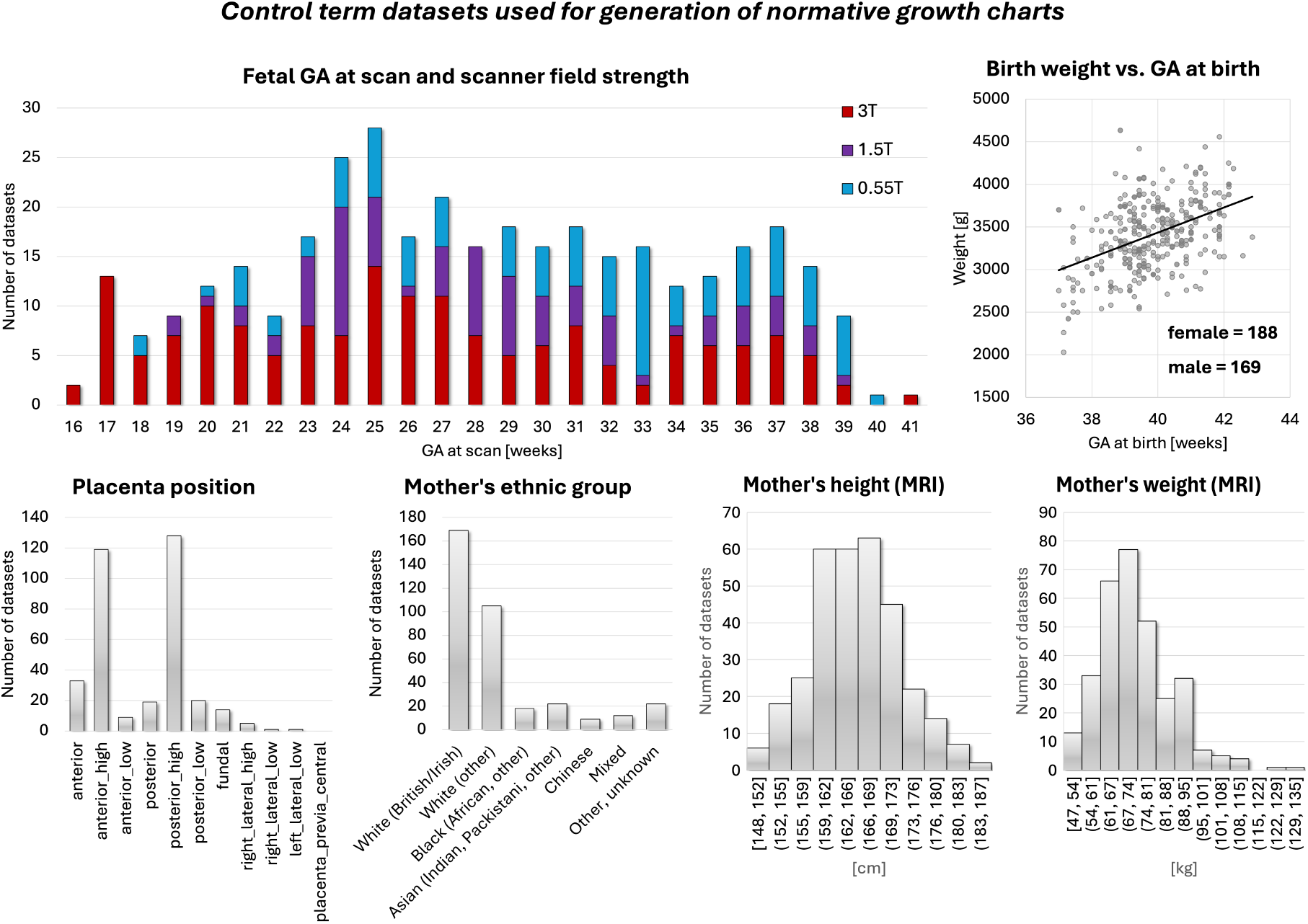
Summary information on the control term cohort (n=357) used for generation of the normative growth charts. *GA* gestational age, *T* Tesla

Fig 5 presents the derived quadratic gestational-age growth curves (5th, 50th, 95th centiles) for fetal, placental and amniotic fluid volumes, as well as the fetal head-to-body volume ratio.

**Fig. 5:**
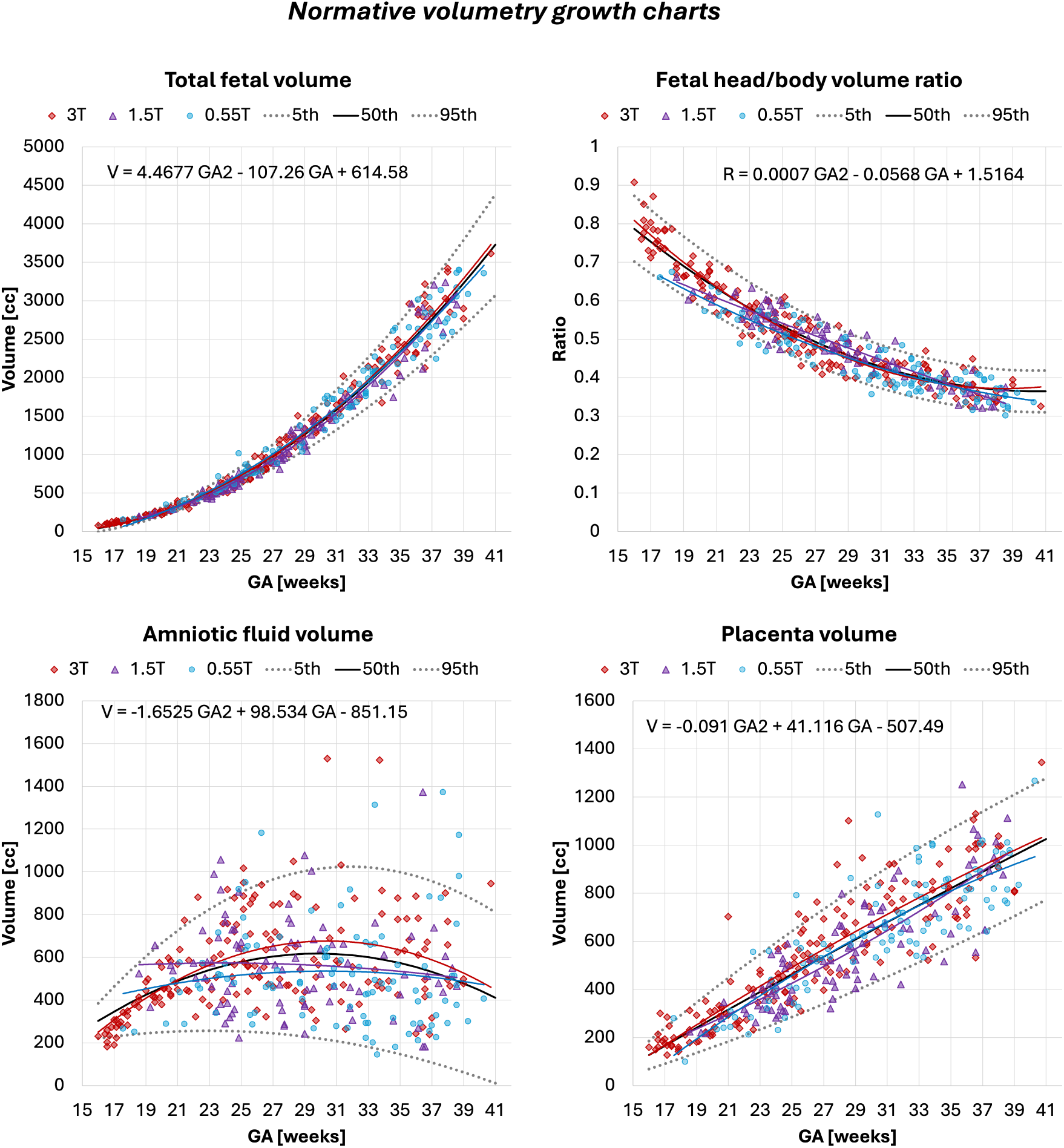
Normative growth charts created from automated volumetry outputs for 357 term control subjects (n=102 at 0.55T, n=88 at 1.5T, n=167 at 3T) with centiles. *GA* gestational age, *T* Tesla

The fetal volume showed the expected strong positive association with gestational age (*p <* 0.0001), with ranges consistent with previously reported MRI norms [12, 15]. Variability increased after approximately 30 weeks, reflecting greater anatomical heterogeneity and expected biological dispersion. The fetal head-body ratio decreased with gestational age (*p <* 0.0001), following known patterns of proportional body growth and relative reduction in head dominance [33].

The placental volume also increased significantly with GA (*p <* 0.0001) within similar ranges to reported earlier works [12, 14]. It showed positive associations with maternal height and maternal weight (both *p <* 0.01) after correction for GA. Placental volume and placental centiles additionally correlated with fetal volume and fetal volume centiles (*p <* 0.0001). This aligns with the well-established relationship between placental mass and fetal growth, as larger placentas typically reflect greater villous surface area and better uteroplacental perfusion [7, 8].

The amniotic fluid volume displayed substantial inter-subject variance across gestation (in agreement with earlier works [12]) but followed a significant quadratic pattern (*p <* 0.0001), with a mid-gestation peak and a decline toward term—consistent with known physiological mechanisms of fetal swallowing, renal output, and placental fluid transfer [34]. Higher amniotic fluid volumes and centiles were positively correlated with placental volume and placental centiles (*p <* 0.01), reflecting the shared dependencies on placental perfusion and fetal health.

After correcting for gestational age at delivery, birth weight was strongly correlated with fetal volume, placental volume, and head-body ratio centiles (*p <* 0.0001). Higher fetal and placental volumes and lower head-body ratios (which reflect proportionately larger fetal bodies) were associated with higher birth weight. Amniotic fluid centiles were also associated with birth weight (*p <* 0.001). Additionally, fetal volume centiles differed between sexes, with higher volumes in male fetuses, matching known sex differences in fetal growth trajectories and birth weight distribution (Fig 6) [35].

**Fig. 6:**
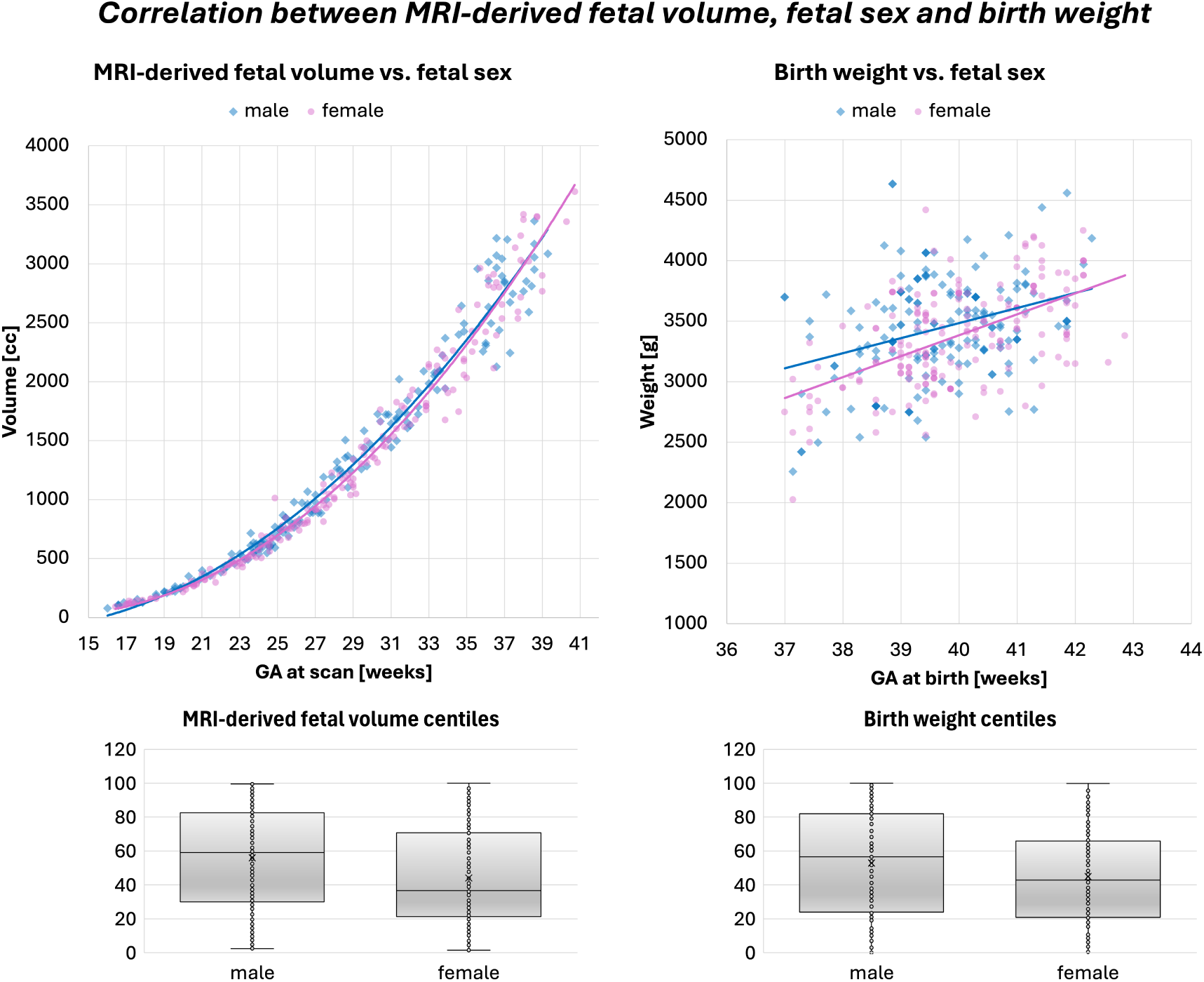
Correlation between MRI-derived fetal volume, fetal sex and birth weight (male - blue, female - pink). *GA* gestational age

Although general volumetric trends overlapped between scanners, ANCOVA revealed significant field-strength effects on fetal volume between 1.5T and 3T (*p <* 0.001) and placenta volume between 1.5T and 3T (*p <* 0.01) with slightly smaller values in 1.5T cohort. These differences are likely driven by unequal GA distributions, a larger proportion of cases at 3T, and other protocol-related factors such as resolution rather than true biological variation. This underlines the importance of harmonised acquisition protocols and careful matching of inclusion criteria when deriving normative volumetric models across multiple field strengths.

To assess longitudinal behaviour, we analysed 95 scans from 42 fetuses, each with two to three scan time points (Fig 7). Fetal volume increased steadily with gestation (146.6 *±* 39.4 cc/week), closely tracking the normative curves. Placental volume also showed consistent growth (38.8 *±* 14.7 cc/week). Amniotic fluid trajectories were more variable (3.3 *±* 35.5 cc/week) but followed the expected pattern of stabilization or decline toward term. Fetal and placental growth rates were only weakly correlated (r *≈* 0.12), whereas fetal growth showed a moderate negative correlation with the amniotic fluid slope (r *≈* 0.44), suggesting that rapid somatic growth often coincided with plateauing or reduced amniotic fluid volume.

**Fig. 7:**
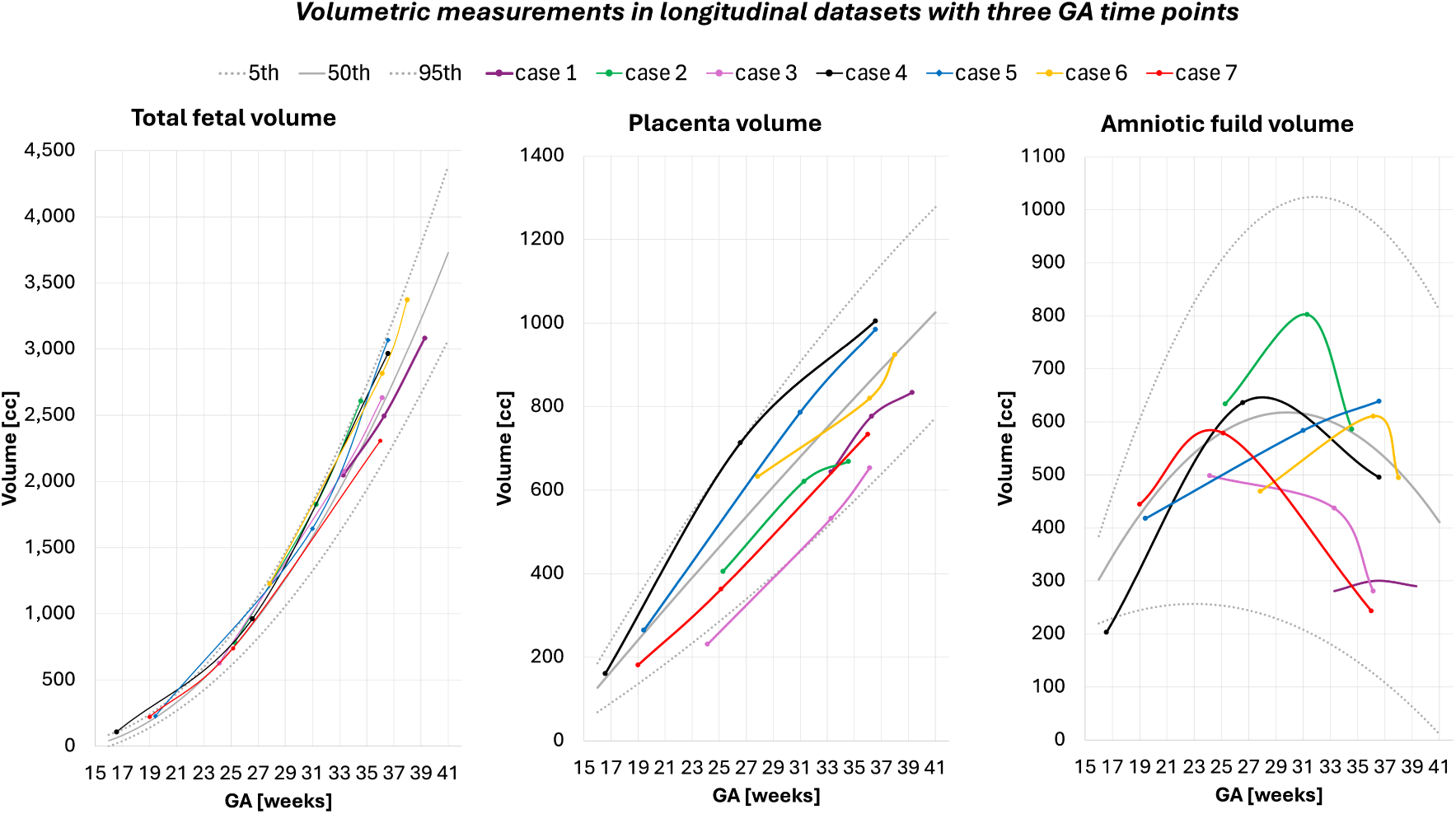
Examples of volumetric changes in longitudinal datasets of 7 subjects scanned at three GA time points. *GA* gestational age

To assess the correlation of patient-specific growth trajectories with the normative models, we performed volumetric analysis of longitudinal datasets of 42 fetuses with multiple GA time points (95 scans in total) from the control term cohort. Fig 7 shows examples of changes in 7 cases with three time points. The fetal volume is demonstrates steady growth with gestation and follows the growth model trend (146.6*±*39.4 cc/week growth rate slope). Placenta volume is also increasing throughout pregnancy similarly to the normative model trajectory (38.8*±*14.7 cc/week growth rate slope). The changes in amniotic fluid volume show high variability between the cases (3.3 *±* 35.5 cc/week growth rate slope) with decreasing trend toward late gestation, which is in agreement with the expected physiological pattern with peak in mid-gestation and then gradually decrease toward term [34]. Across fetuses, the individual growth rates of fetal and placental volume were only weakly correlated (*r ≈* 0.12). Interestingly, fetal growth rate showed a moderate negative correlation with the amniotic fluid slope (*r ≈* 0.44), suggesting that cases with rapidly increasing fetal volume tended to show stable or declining amniotic fluid volumes

Overall, these findings demonstrate the validity and clinical relevance of the generated normative volumetry models, while highlighting the need for future work incorporating detailed demographic factors, abnormal cases, and outlier centiles to support personalised growth prediction and improved diagnostic stratification.

### Volumetric analysis of term vs. preterm cohorts

To assess the feasibility and clinical utility of the proposed automated volumetry pipeline, we applied it to a heterogeneous abnormal cohort of 86 preterm datasets that included a mix of pregnancies with PPROM and pregnancies with intact membranes, all resulting in delivery *≤*32 weeks GA, and compared these with 252 term-born controls scanned at *≤*32 weeks GA. The preterm group (Fig 8a) encompassed a range of clinical indications and pregnancy complications, including spontaneous preterm birth, preeclampsia requiring elective delivery, and PPROM, as well as PPROM cases electing for termination. These participants were recruited on the basis of a high predicted risk of preterm birth and subsequently delivered at < 32 weeks GA.

**Fig. 8:**
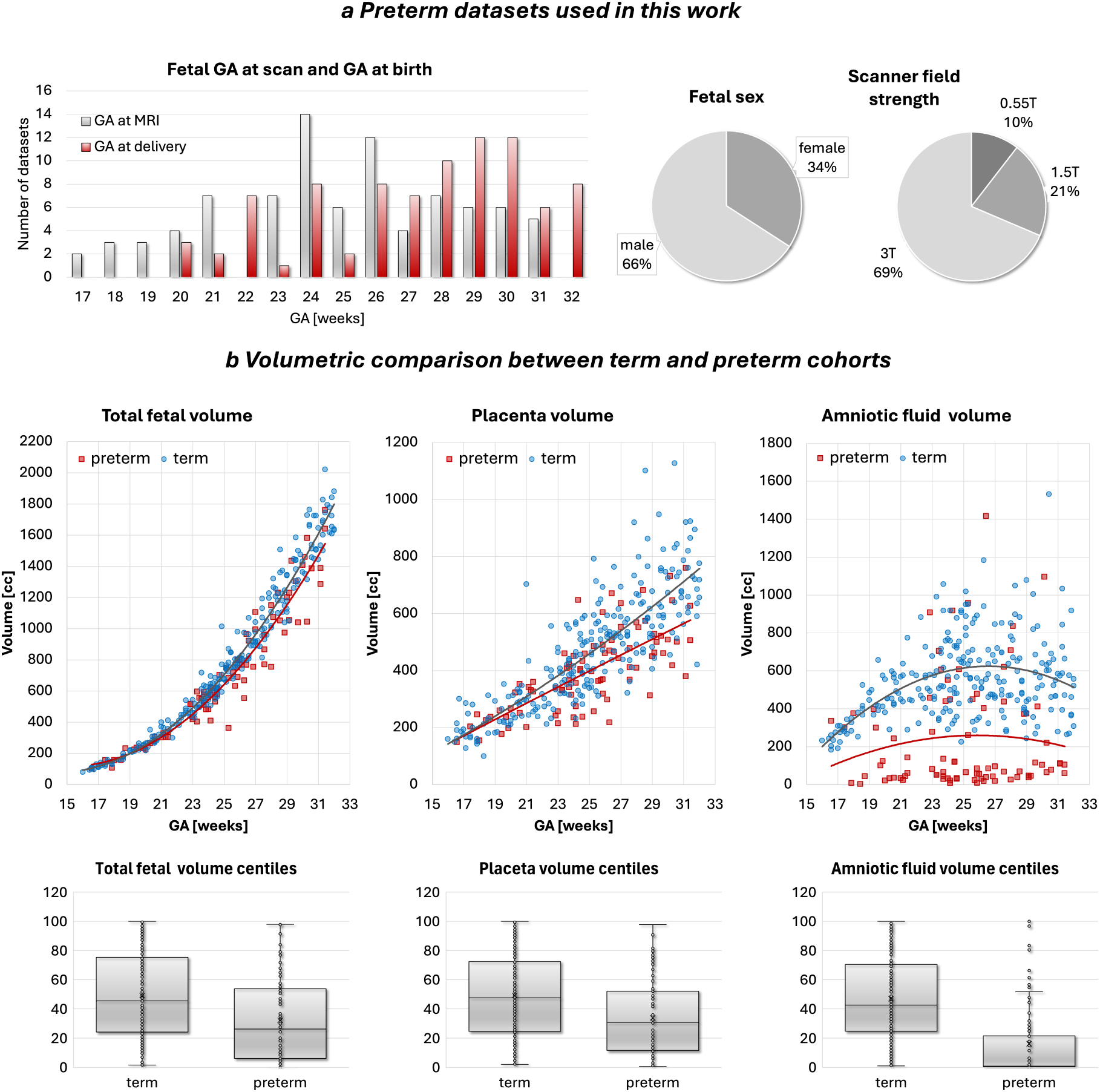
Term vs. preterm cohort analysis. a Summary information on the abnormal preterm cohort (n=86) used in feasibility sub-study. b Volumetric comparison between term (n=252) and preterm (n=86) cohorts. *GA* gestational age, *MRI* magnetic resonance imaging, *T* tesla

All preterm datasets were segmented automatically, followed by visual review. Manual refinement was required in 35 cases, mainly in the placenta and fetal body ROIs. This was typically due to reduced soft-tissue visibility and marked amniotic fluid loss in PPROM, and refinements were brief (< 5 minutes per case). These findings highlight the importance of further model retraining for atypical anatomy and underline the need for automated quality-control methods within future versions of the pipeline.

As shown in Fig 8b, fetal volumes in the preterm cohort show a broad spread but a significantly lower trendline compared with the term controls (*p <* 0.0001), reflected in lower fetal volume centiles (*p <* 0.0001) in agreement with earlier studies [36]. Placental volumes and placental centiles were likewise markedly lower in preterm pregnancies (*p <* 0.0001), consistent with impaired placental development or compromised uteroplacental perfusion described in preterm birth and hypertensive disorders [7].

Amniotic fluid volumes in the preterm cohort were also significantly reduced, particularly in PPROM cases, where membrane rupture results in fluid loss. This trend was reflected in both absolute volume (*p <* 0.01) and centiles (*p <* 0.0001).

Examples of automatically generated .html volumetry reports for PPROM cases are shown in Fig 9. In both examples, amniotic fluid centiles were < 5*th* centile. In one case, both fetal and placental volumes were < 5*th* centile, while in the other case, fetal and placental volumes remained within normal ranges despite severe oligohydramnios. The report format and centile-based interpretation were reviewed by fetal MRI clinicians and considered clear, clinically meaningful, and useful for integration into routine reporting workflows.

**Fig. 9:**
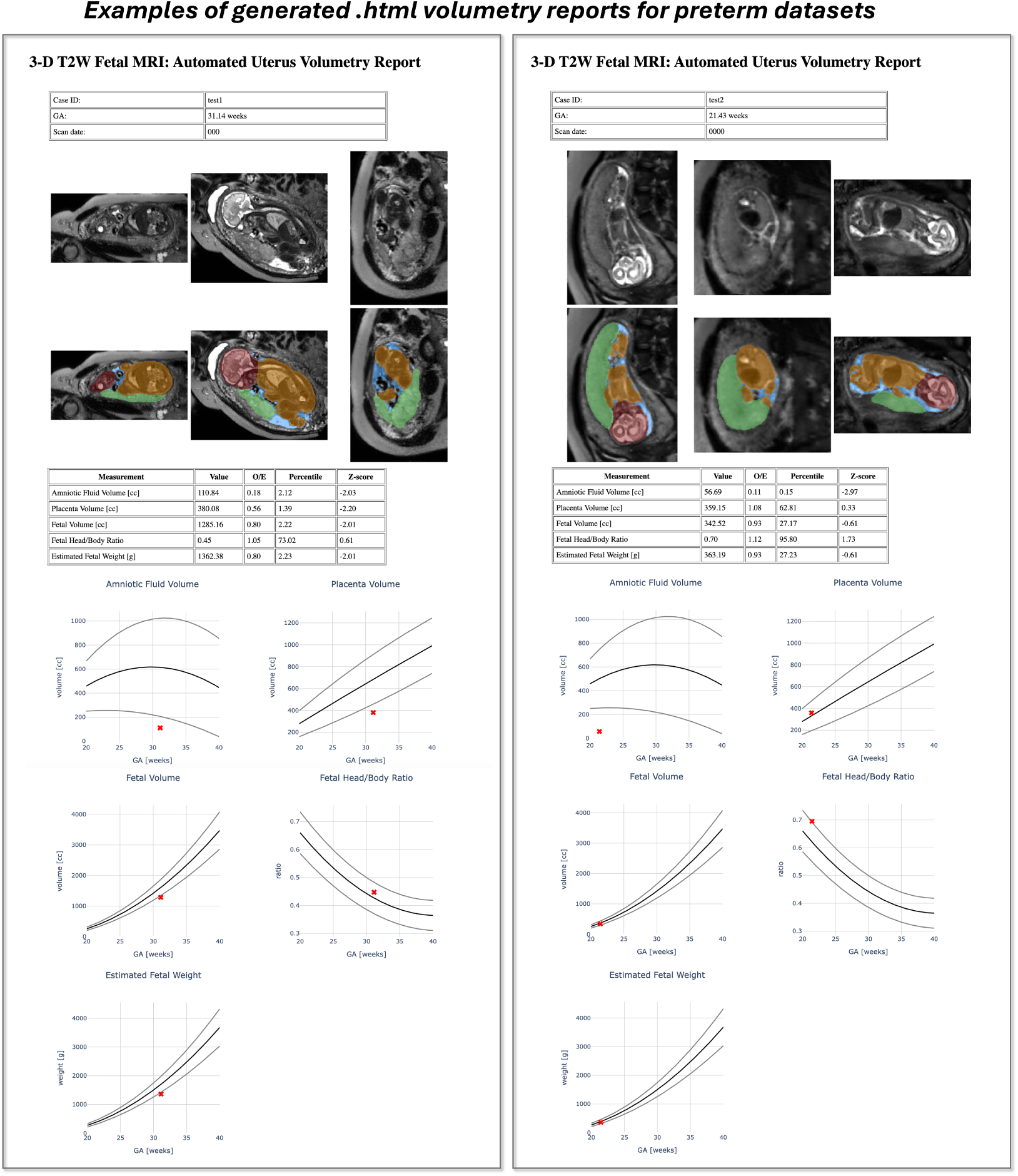
Examples of automated reports generated as .html files. *3-D* 3-dimensional, *GA* gestational age, *MRI* magnetic resonance imaging, *O/E* observed-to-expected, *T2W* T2-weighted

Together, these results demonstrate that the automated volumetry pipeline is capable of capturing meaningful physiological differences between normal and highrisk pregnancies, and support the feasibility of its application in clinical and research settings. They also illustrate the potential for this tool to serve as a baseline framework for further refinement and retraining toward specific clinical scenarios and pathology-focused models.

## Discussion

This study presents the first integrated framework for automated volumetry of the fetus, placenta and amniotic fluid in motion-corrected 3-D reconstructed T2W fetal MRI across 0.55T, 1.5T and 3T acquisition protocols. Using deformable slice-to-volume reconstruction and a 3-D UNet, we derived normative volumetric growth charts from a large and well-defined cohort of 357 confirmed healthy term-control pregnancies scanned between 16-41 weeks GA. This directly addresses longstanding limitations of prior MRI volumetry, which relied on labour-intensive manual segmentation of 2-D motion-corrupted stacks with DL models predominantly trained to segment only a single ROI. Importantly, there is also lack of combined true normative MRI volumetry models for the fetus, placenta or amniotic fluid generated from confirmed healthy control cohorts, despite the clinical need for reliable reference curves for z-scores and abnormality detection.

The segmentation model performed robustly across field strengths and gestational ages, with only minor manual refinements required—mostly at the placenta-myometrium interface, a known area of low contrast in fetal MRI. Performance across heterogeneous acquisition protocols supports the generalisability of the approach and its utility in both retrospective and prospective datasets.

The derived growth curves reproduced well-established physiological patterns. Fetal volume increased steadily with widening variance toward term, consistent with ultrasound and MRI literature [15]. Placental volume increased throughout gestation, reflecting villous maturation and vascular expansion, and showed positive associations with maternal height, weight and birth weight—relationships that align with known correlations between placental mass, uteroplacental capacity and neonatal size. Amniotic fluid volumes displayed the expected high inter-subject variability and characteristic mid-gestation peak with a decline toward late pregnancy, reflecting the changing balance between fetal urine production, swallowing and intramembranous absorption.

MRI-derived volumetry also demonstrated meaningful correlations with birth weight. Higher fetal and placental volumes, and lower fetal head-body ratios, were associated with higher birth weight after GA correction. Sex-related differences in total fetal volume (larger in males) further matched established biological patterns.

Analysis of 95 longitudinal scans from 42 term-control fetuses showed that fetal and placental growth trajectories followed the normative curves, whereas amniotic fluid slopes were more variable—consistent with the dynamic physiology of amniotic fluid regulation. These findings highlight the potential of MRI volumetry for personalised growth assessment when ultrasound is inconclusive or insufficient.

Application to 86 preterm pregnancies delivered at ≤32 weeks demonstrated significantly lower fetal, placental and amniotic fluid volumes and centiles relative to matched term controls. These findings align with known impairments in placental function, fetal growth and membrane integrity in PPROM, preeclampsia and spontaneous preterm birth. Mild refinements were occasionally required in cases with severe fluid loss, but automated segmentation remained feasible across the cohort, indicating real-world clinical applicability.

Overall, our findings demonstrate the value of automated whole-uterus volumetry for detecting deviations from expected growth trajectories in both cross-sectional and longitudinal settings. This pipeline provides a practical foundation for quantitative fetal and placental MRI.

### Limitations and future work

In terms of limitations, segmentation accuracy remains partly dependent on DSVR reconstruction quality and local image contrast. The placenta-myometrium interface and fetal extremities continue to present challenges, particularly in PPROM cases with markedly reduced amniotic fluid. Future work could incorporate targeted augmentation, contrast-aware architectures or attention-based models to reduce the need for manual refinement, alongside automated quality-control scoring to improve reliability. The normative models were derived from healthy pregnancies with confirmed delivery at term, but several relevant biological and demographic modifiers—such as fetal sex, maternal age, height, weight, ethnicity and general health or fitness—were not explicitly incorporated, despite their known influence on fetal and placental size. Future extensions will focus on demographic-adjusted normative curves and mixed-effects or Gaussian-process-based modelling, which may better accommodate the nonlinearity of growth trajectories. Broader inclusion of ethnically and socioeconomically diverse cohorts will also be essential for improving geographic and population-level generalisability.

The current pipeline focuses on global intrauterine structures. Extending the framework to additional fetal organs (e.g., lungs, liver, brain) [37, 38] would support organ-specific volumetry pipelines, which frequently rely on total fetal volume for normalisation. Further integration of placental surface area estimation, automated placental site characterisation, umbilical cord insertion localisation, and fetal lie assessment could enrich the utility of whole-uterus analysis.

Finally, multi-centre validation across vendors, field strengths and acquisition protocols will be important to establish generalisability and accelerate adoption in routine clinical workflows. Complementary studies incorporating paired MRI-ultrasound volumetry, both for whole-uterus and organ-specific measurements, will offer insight into cross-modality reproducibility and help define how automated MRI volumetry can best complement established ultrasound-based assessment.

## Conclusion

We present the first automated framework for simultaneous fetal, placental and amniotic fluid volumetry in motion-corrected 3-D reconstructed T2-weighted fetal MRI, together with the first combined normative volumetric growth models derived from a large cohort of confirmed healthy term-control pregnancies at 0.55-3T scanner field strengths. The pipeline provides efficient, standardised volumetric assessment across a wide range of MRI field strengths, supports individualised interpretation through z-scores and centiles and performs reliably in both normal and high-risk pregnancies. Demonstrated applicability in longitudinal and preterm cohorts underscores its potential for integration into routine fetal MRI assessment and future quantitative fetal and placental research.

## Acknowledgments

We thank everyone who was involved in acquisition and analysis of the datasets at the Department of Perinatal Imaging and Health at Kings College London and St Thomas’ Hospital. We thank all participants and their families.

This work was supported by the NIHR Advanced Fellowship to L.S. [NIHR3016640], the MRC grant [MR/W019469/1], the MRC grant [MR/X010007/1], the Wellcome Trust, Sir Henry Wellcome Fellowship to J.H. [201374/Z/16/Z], the UKRI FLF to J.H. [MR/T018119/1], DFG Heisenberg [502024488] the High Tech Agenda Bavaria to J.H., the NIH Human Placenta Project grant [1U01HD087202-01], the Wellcome/EPSRC Centre [WT203148/Z/16/Z], the NIHR Clinical Research Facility (CRF) at Guy’s and St Thomas’ and by the National Institute for Health Research Biomedical Research Centre based at Guy’s and St Thomas’ NHS Foundation Trust and King’s College London.

The views expressed are those of the authors and not necessarily those of the NHS, the NIHR or the Department of Health.

## Data availability

The individual fetal MRI datasets used for this study are not publicly available due to ethics regulations. For more information please contact Jana Hutter.

## Author contributions

AU developed deep learning segmentation pipeline and reporting script, processed the datasets, performed testing and analysis, and prepared the manuscript. MH contributed to formalisation of the whole fetus, amniotic fluid and placenta parcellation protocol and performed manual segmentations. CB, AK participated in analysis and acquisition of the datasets. JAV, SNS, KC contributed to acquisition of the datasets. HW, AEC, PAG, LCG, AL, VK, MD, JM contributed to analysis of the datasets. JVH, MR and JH provided fetal MRI datasets and supervised various parts of the project. LS provided fetal MRI datasets, contributed to formalisation of the parcellation protocol, performed manual segmentations, and evaluation and supervised all stages of the project. All authors reviewed the manuscript.

